# Reduced Neural Speech Tracking in Adolescents with Listening Difficulty

**DOI:** 10.1101/2025.06.24.25330187

**Authors:** Katsuaki Kojima, Chunyan Liu, Shelley Ehrlich, Harvey Dillon, Chelsea Blankenship, Lina Motlagh Zadeh, Jennifer Vannest, Lisa Hunter, Srikantan Nagarajan, David R. Moore

## Abstract

**Objective:** To investigate neural mechanisms underlying speech-in-speech listening in adolescents with listening difficulties (LiD).

**Methods:** Neural speech tracking (NST) was assessed using magnetoencephalography (MEG) in 21 adolescents with LiD and 25 typically developing (TD) peers, all with audiometrically normal hearing. Participants performed a cocktail party task involving target speech presented alone or alongside competitor speech streams differing in talker identity and spatial location. NST was quantified using theta-band (4–8 Hz) inter-event phase coherence (IEPC) to acoustic edges.

**Results:** Adolescents with LiD demonstrated significantly reduced NST of target speech relative to TD peers, whereas NST of competitor speech was comparable between groups. Theta-band IEPC correlated with caregiver-reported listening difficulties in both groups, indicating clinical relevance. The presence of both talker and spatial cues synergistically enhanced NST of target speech, an effect more pronounced in TD adolescents and absent for competitor streams.

**Conclusions:** LiD is characterized by impairment in processing attended speech rather than enhanced competitor processing or generalized auditory disengagement. Reduced cue integration observed in adolescents with LiD may contribute to their listening challenges.

**Significance:** These findings suggest targeted therapeutic interventions enhancing selective auditory attention and multimodal cue integration may effectively address listening difficulties in adolescents with LiD.

**Highlights:**

- Adolescents with listening difficulty showed selective speech-tracking deficits in attended speech.
- Spatial and talker cues synergistically enhance neural tracking of attended speech.
- Neural speech tracking measures correlate with caregiver-reported listening difficulties.

## 1. INTRODUCTION

Listening difficulty (LiD) is a condition linked to developmental language disorders and characterized by challenges in understanding speech, including in complex listening environments such as when multiple people are talking simultaneously and from different directions (Dillon and Cameron 2021; Dawes and Bishop 2009). This listening situation has been called the cocktail party problem (Cherry 1953) or competing speech listening. Children with LiD frequently struggle to follow conversations and directions in noisy classrooms or social settings, which can lead to persistent challenges in communication, cognitive function, and psychosocial health (Del Zoppo et al. 2015; Lawton et al. 2017). Despite having clinically normal audiometric thresholds, children with LiD exhibit deficits in more complex auditory skills, such as auditory discrimination, dichotic listening, and temporal processing (Rosen et al. 2010; Iliadou et al. 2010; Phillips et al. 2010). Through a longitudinal cohort study, Sensitive Indicators of Childhood Listening Difficulties (SICLiD), our research group has investigated the mechanisms underlying LiD. This study has revealed significant differences between children with LiD and typically developing (TD) controls across multiple domains, spanning speech-in-speech test performance, neural responses to amplitude modulation, cognitive performance, functional brain connectivity, and socioeconomic factors (Petley et al. 2021; Stewart et al. 2022; Petley et al. 2024; Kojima et al. 2024).

However, despite these advances, neural mechanisms underlying speech processing differences between LiD and TD individuals during competing speech listening remain poorly understood. A critical gap in LiD research is the limited understanding of the distinct neural processing of target and competitor speech, as previous studies have predominantly focused on target speech processing alone (Krishnamurti 2001; Mattsson et al. 2019). Research beyond the LiD domain has employed both invasive and non-invasive neuroimaging methods in adults to investigate cortical representations of concurrent speech streams (Ding and Simon 2012; Mesgarani and Chang 2012; Zion Golumbic et al. 2013).

Electrocorticography studies have revealed neural encoding of separate target and competitor streams, with attenuated representation of competitor speech streams in the auditory cortex, highlighting the modulatory role of selective attention to target streams (Mesgarani and Chang 2012; Zion Golumbic et al. 2013). Complementary magnetoencephalography (MEG) findings have shown distinct encoding of target and competitor speech streams, with enhanced fidelity observed for target streams (Puvvada and Simon 2017).

Extending these insights to LiD populations allows testing three mechanisms that might explain differences in competing speech processing between TD and LiD groups. First, children with LiD may exhibit heightened neural responses to competitor speech stream alongside typical tracking of the target stream, suggesting impaired suppression of irrelevant auditory inputs, as indicated by prior behavioral and neurophysiological evidence in attention-related auditory deficits (Hillyard et al. 1973; Bharadwaj et al. 2015; Getzmann and Falkenstein 2011). Alternatively, individuals with LiD might show diminished neural responses to both target and competitor speech streams, indicative of a generalized auditory processing deficit consistent with findings of reduced cortical auditory processing efficiency observed in language disorders (Sharma et al. 2009; Bishop et al. 2012). Third, LiD could involve typical neural encoding of competitor speech but impaired neural tracking of the target stream, aligning with evidence of selective attention or cognitive load impairments during auditory stream segregation tasks observed clinically in individuals without broader auditory or attentional deficits (Anderson et al. 2013; Oberfeld and Klockner-Nowotny 2016).

Unlike the first mechanism—primarily indicative of a broad attentional control deficit—and the second— suggesting a generalized auditory-language impairment—the third mechanism aligns most closely with the hallmark clinical presentation of LiD: difficulty tracking target speech amid competing talkers. This conceptual fit is reinforced by our enrollment strategy, which relied on the Evaluation of Children’s Listening and Processing Skills (ECLiPS), a comprehensive caregiver-rated instrument that quantifies real-world listening challenges in multi-speaker environments (Barry et al. 2015; Roebuck and Barry 2018; Denys et al. 2024; Barry and Moore 2021). Neurophysiological evidence further supports this view: increasing the linguistic load of a distractor markedly suppresses cortical entrainment to the attended stream without boosting neural encoding of the distractor itself, indicating that interference chiefly arises from degradation of the target representation rather than from abnormal capture of neural resources (Dai et al. 2022). Together, the consonance between the clinical phenotype of LiD, our participant selection criteria, and prior electrophysiological findings leads us to hypothesize that children with LiD will exhibit intact encoding of competing speech but reduced neural tracking of the target stream during competing-speech listening.

Neural speech tracking (NST), a method quantifying neural phase-locking responses to speech envelope fluctuations, is particularly well-suited for investigating neural encoding of target and competitor speech streams simultaneously during competing speech listening tasks (Ding and Simon 2012; Puvvada and Simon 2017). The speech envelope—characterized by slow amplitude modulations—carries essential linguistic information such as syllabic structure, prosody, and stress patterns, which aid listeners in differentiating target speech from competitors (Drullman et al. 1994; Goswami et al. 2010; Myers et al. 2019; Oganian and Chang 2019; Swaminathan et al. 2016). Crucially, neural phase-locking to this envelope synchronizes neural excitability with prominent acoustic features, particularly acoustic edges—rapid amplitude increases typically marking syllable onsets (Doelling et al. 2014; Oganian and Chang 2019). These acoustic edges serve as temporal anchors, facilitating segmentation of continuous speech into linguistically meaningful units. Our previous work combining simulations and MEG recordings from healthy adults during natural speech listening demonstrated that NST predominantly reflects neural phase-locking to acoustic edges within the theta frequency band (4–8 Hz) (Oganian et al. 2023). Given this robust foundation, we will employ this NST approach to elucidate the differential neural processing of target and competitor speech streams in children with LiD and TD peers.

During competing speech listening, listeners can leverage multiple acoustic cues, such as talker identity and spatial location information, to distinguish target speech streams from competitors (Kidd et al. 2010; Bronkhorst 2015). Talker cues emerge from distinct spectrotemporal properties of individual voices, including fundamental frequency, vocal tract length, and speaking rate, which become available when target and competitor streams are produced by different speakers (Darwin et al. 2003; Newman and Sawusch 2009). Spatial cues arise from the differential positioning of speech sources in the acoustic environment, enabling listeners to separate speech streams through binaural processing of interaural time and level differences—variations in sound arrival time and intensity between the ears that provide crucial spatial information (Bronkhorst and Plomp 1988; Litovsky 2012). While both cue types facilitate speech stream segregation, their interactive effects and underlying neural mechanisms to utilize these cues remain subjects of ongoing investigation. Some studies suggest these cues operate through parallel and independent processing pathways (Du et al. 2011), while others indicate potential interference or competitive integration between the cue types (Wang et al. 2021; Allen et al. 2011). This complexity is particularly relevant in understanding LiD, as behavioral studies have demonstrated that children with LiD exhibit reduced ability to utilize both talker and spatial cues compared to TD children (Cameron et al. 2006b; Flanagan et al. 2018; Petley et al. 2021; Kojima et al. 2024). However, the neural mechanisms underlying this impairment remain poorly understood, particularly regarding how individuals with LiD integrate multiple acoustic cues during speech processing and how these cues modulate NST in complex listening environments.

The present study leverages our longitudinal cohort of adolescents with LiD and TD controls to explore the neural mechanisms underlying listening difficulties during competing speech listening tasks. Using MEG recordings, we quantify NST through phase-locking responses to acoustic edges in the theta band during a cocktail party task. To test mechanistic accounts, we evaluate two specific hypotheses: First, we predict that adolescents with LiD will demonstrate reduced NST of target speech compared to TD adolescents, while maintaining similar tracking of competitor—a pattern that would suggest selective deficits in processing attended speech. Second, we hypothesize that while both talker and spatial cues will enhance neural tracking of target speech in TD adolescents, this enhancement will be significantly reduced in adolescents with LiD, indicating impaired utilization of these crucial cues for speech stream segregation.

## 2. METHODS

### 2.1. Participants

Participants were recruited from the parent study (SICLiD) and from employee families. All participants were aged 10–19 years, had English as their primary language, and demonstrated clinically normal hearing thresholds (≤ 20 dB HL, bilaterally; 0.25–8 kHz). Exclusion criteria for both the TD and LiD groups included the presence of neurologic, psychiatric, or intellectual conditions that could interfere with their ability to complete the study procedures. Participants in the TD group had no reported history of listening difficulties, cognitive impairments, or learning disorders.

Assignment to the LiD or TD groups was based on performance on the ECLiPS Total Scaled score. Participants scoring below the 10th percentile, or a numerical score of 6, on the ECLiPS were assigned to the LiD group while the remaining children were assigned to the TD group (Petley et al. 2021; Kojima et al. 2024). Groups were frequency-matched for age, sex, race, and ethnicity.

Initially, 22 LiD and 16 TD participants were recruited from the SICLiD study, with an additional 12 TD participants recruited from Cincinnati Children’s Hospital employee families. Three participants (1 LiD and 2 TD) were excluded due to abnormal audiometry results, and one TD participant did not complete the data collection. The final sample included 21 LiD and 25 TD participants.

To maintain the integrity of the group classifications, participants with a history of LiD (as defined by prior ECLiPS scores below the 10th percentile during earlier SICLiD visits) were retained in the LiD group, even if their ECLiPS scores at the time of the current study visit exceeded the 10th percentile. Conversely, participants previously classified as TD in the SICLiD study who scored below the 10th percentile during the current study visit were reclassified into the LiD group. This approach ensured that the LiD group included individuals with even subtle risks of listening difficulties, while the TD group comprised only healthy controls.

The study was approved by the Institutional Review Board of Cincinnati Children’s Hospital Medical Center. Written informed consent was obtained from parents or legal guardians of participants under 18 years of age, while participants aged 18 years or older provided consent for themselves. All participants received financial compensation for their time and participation.

### 2.2. Speech Stimuli

The experimental stimuli comprised five 5-minute stories from a children’s news podcast (KidNuz.org), narrated by a female talker (“Tori”) (for full stimulus transcripts, see Supplemental Table S2). Four conditions were competing speech listening, where participants heard one target speech stream simultaneously with two competitor streams, while the fifth was a “clear” condition without competitors (Figure 1B). To facilitate clear MEG onset response measurements, the target streams contained 500-700 ms silent intervals between sentences.

**Figure 1.**
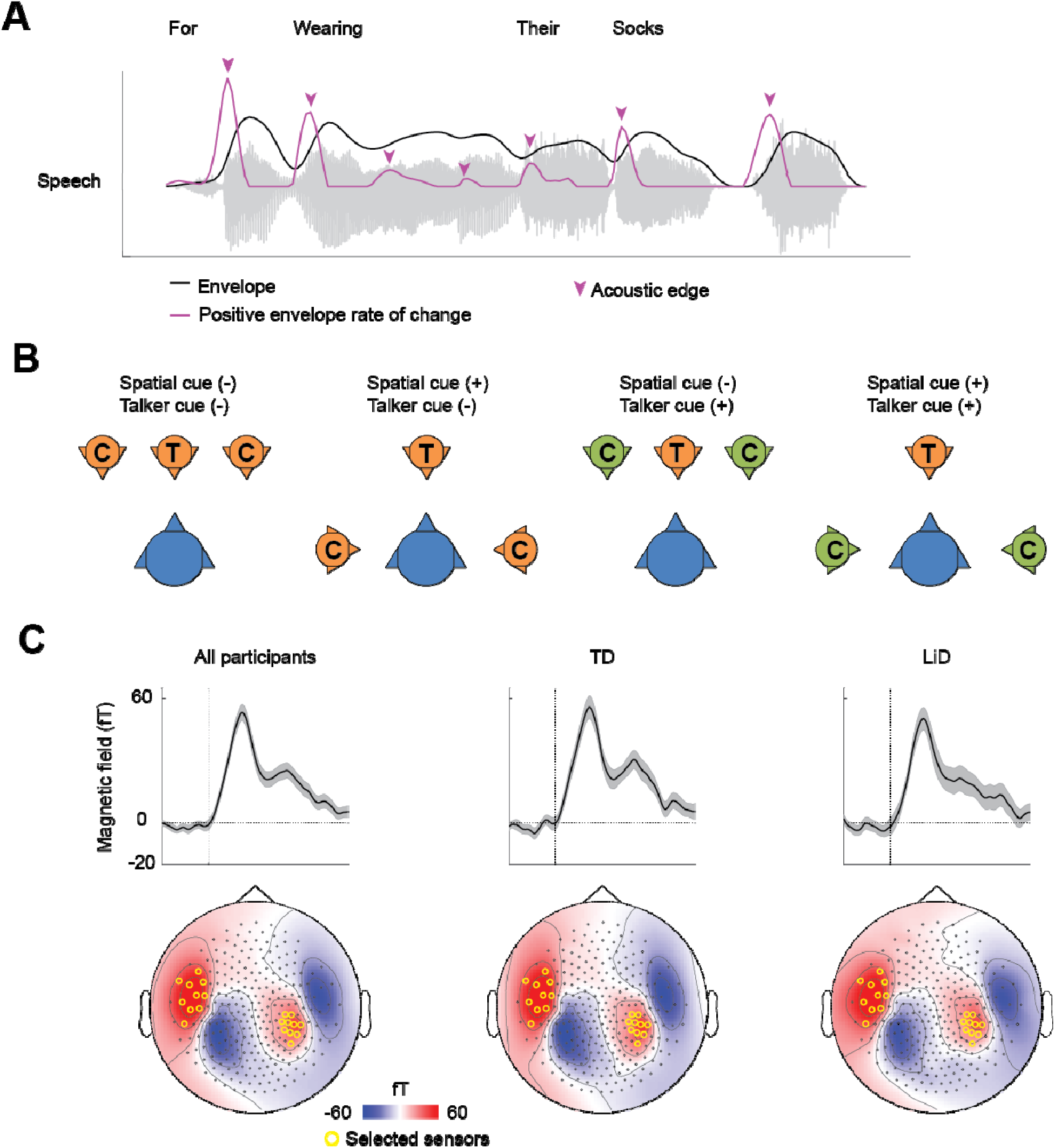
Task design and auditory sensor selection based on M100 responses. **A.** The acoustic waveform of an example utterance (“For wearing their socks”) displayed with its amplitude envelope (black line) and corresponding first temporal derivative (purple line). Arrows indicate acoustic edges, defined as local peaks in the derivative of the envelope. **B.** Schematic representation of virtual spatial configurations and talker identity conditions for target (T) stream and competitor (C) across four competing speech conditions. Spatial cues are present when the target and competitor have different virtual positions (2nd and 4th panels). Talker cues occur when the target and competitor have different voices (3rd and 4th panels). **C.** Sensor selection (highlighted in yellow) was determined by M100 responses to sentence onsets. Upper panels show evoked responses, lower panels display M100 topographies for the total participant cohort (left), TD (middle), and individuals with LiD (right). No significant differences in evoked responses were observed between TD and LiD groups (p > .05, cluster-based permutation testing).

The competing speech conditions were systematically varied across two dimensions: spatial and talker cues (Figure 1B). Spatial differentiation was achieved by virtually positioning the competitor talkers either at 90 degrees to the right and left of the target talker (spatial cue present) or at the same angle as the target talker (spatial cue absent). Virtual talker locations were implemented using binaural head-related transfer functions (HRTFs) recorded in an anechoic chamber above 50 Hz using a Knowles Electronics Manikin for Acoustic Research with small pinnae (Cameron et al. 2006a). Talker cue was manipulated by using either a different female talker (“Kim”) or the same talker (“Tori”) for the competitor streams.

Acoustic analysis revealed distinct characteristics between talkers. The target talker (Tori) exhibited a lower mean fundamental frequency (197 Hz, SD = 1.9 across listening conditions) compared to the competing talker (Kim: 204 Hz, SD = 2.0 across listening conditions; t(6.0) = −5.2, p = .002). While both talkers showed substantial within-utterance F0 variability (typical SD within conditions = 65-71 Hz), their mean F0 values remained consistently different across conditions. Additionally, Tori’s speech rate, measured by acoustic edge frequency, was higher (6.8 Hz, SD = 0.2) than Kim’s (6.5 Hz, SD = 0.1; t = 3.4, p = .01). Detailed acoustic characteristics are provided in the supplemental Table S1 and Figure S1.

The target streams during clear and competing speech conditions were calibrated to −25 dB root mean square amplitude, while competitors were set 5 dB lower than the target speech streams. This signal-to-noise ratio (SNR) was determined through pilot recordings to optimize the balance between maintaining task feasibility and eliciting measurable differences in IEPC. Previous research using multiple SNRs has demonstrated that this level enables reliable speech tracking while remaining sensitive to differences between LiD and TD participants (Vander Ghinst et al. 2021; Hausfeld et al. 2021).

### 2.3. Procedure and stimulus presentation

Binaural auditory stimuli were presented to participants at a comfortable ambient loudness (∼70dB) through an MEG-compatible audio system using Presentation® software (Neurobehavioral Systems, Albany, CA, USA). The audio system consisted of a distal transducer connected via acoustic tubing to ear inserts (Etymotic Research, IL, USA), which delivered auditory stimuli directly to participants’ ears. Speech stimuli were digitized at a sampling rate of 44.1 kHz. Participants were instructed to listen attentively to the target speech throughout the experiment.

The experiment consisted of five 5-minute listening blocks: one clear speech condition and four competing speech conditions. All participants completed the clear speech condition first, followed by the four competing speech conditions. The order of the competing speech conditions was counterbalanced across participants to minimize order effects, with participants randomly assigned to one of two possible orders. After each listening block, participants answered seven multiple-choice questions assessing their comprehension of the target speech content (for a list of comprehension questions, see Supplemental Table S3).

### 2.4. Neural data acquisition

MEG data were acquired using a 275-channel whole-head CTF system (MEG International Services Ltd., Coquitlam, BC, Canada) with real-time noise cancellation achieved through third-order gradiometer formation. The system has very low intrinsic noise per superconducting quantum interference device (SQUID) detector. Neuromagnetic activity was recorded at a sampling rate of 1200 Hz while participants listened to acoustic stimuli (details in section 2.2) in a supine position. Head localization coils were placed at the nasion and preauricular locations to ensure accurate localization of neural activity.

Continuous head localization was performed at 30 Hz throughout the recording session. To minimize the effects of head movement on data quality, only recordings with less than 5 mm of total head movement were accepted for further analysis.

### 2.5. Neural data analysis

All analyses were conducted in MATLAB R2023b (The MathWorks, https://www.mathworks.com) using custom-written scripts and the FieldTrip toolbox (Oostenveld et al. 2011). The analysis steps were similar to our prior published work (Oganian et al. 2023), including acoustic feature extraction, sensor selection, and calculating IEPC.

#### 2.5.1. Acoustic feature extraction

The broad amplitude envelope of the speech stimuli was calculated by rectifying, low-pass filtering at 10 Hz, and downsampling to 100 Hz the original stimulus waveform. The derivative of the resulting envelopes was then computed to measure its rate of change. Local peaks in the derivative of the amplitude envelope (acoustic edges) were extracted as a sparse time series (Oganian and Chang 2019). Figure 1A depicts these features for an example excerpt. For competing speech conditions, acoustic edges were extracted separately for the target stream and competitors using the audio data prior to mixing. The number of acoustic edges was higher in the audio files read by Tori compared to Kim (Tori: M = 1810, SD = 71; Kim: M = 1670, SD = 56; t = 3.8, p = .006).

#### 2.5.2. MEG data preprocessing

MEG data were preprocessed offline using artifact rejection with dual signal subspace projection (DSSP) (Sekihara et al. 2016) and downsampled to 400 Hz. DSSP, a validated MEG interference rejection algorithm based on spatial and temporal subspaces, effectively removes artifacts from clinical data (Cai et al. 2019). In subsequent analyses, segments containing single sensor data exceeding 1.5 pT or visually identified artifacts (muscle, eye blink, and motion) were removed as bad events (0.3% of segments).

#### 2.5.3. Sensor selection

To focus on responses originating in temporal auditory areas, sensors were selected based on the magnitude of the group-averaged M100 response to sentence onsets. The broadband signal was segmented around sentence onsets (−200 to 500 ms), averaged across sentences and participants, and baseline corrected (−200 ms to 0 ms). M100 amplitude was extracted as the average activity between 50 and 100 ms after sentence onset. The ten sensors with maximal M100 responses per hemisphere (20 total) were selected for subsequent analyses.

#### 2.5.4. Time-frequency (TF) decomposition

The instantaneous phase of the neural signal was calculated at individual frequency bands, logarithmically spaced between 0.2 and 10 Hz in 1/7 octave steps. Noncausal bandpass Butterworth filters were applied around each frequency of interest with a filter width of ± 0.1 octaves. The filter order was chosen to achieve a maximum of 3 dB passband ripple and at least 24 dB stopband attenuation. The Hilbert transform was then performed on the filtered signals. The amplitude and phase were obtained as the absolute value and phase angle, respectively, of the Hilbert signal.

#### 2.5.5. Inter-event phase coherence (IEPC)

We assessed neural phase-locking around acoustic edges by segmenting the continuous phase data around these events and calculating a time-resolved IEPC (Lachaux et al. 1999). IEPC was computed for each time point using the following formula:

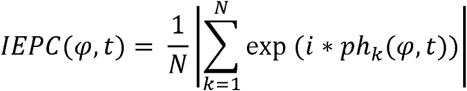

where N is the number of events, ph is the phase of the neural signal in trial k, for frequency band w and time point t. IEPC was first calculated within each selected sensor, then averaged across sensors.

To assess the distribution of phase-locking following acoustic edges, we segmented the phase data output by the TF analysis around acoustic edges (−500 to 500 ms) and calculated the IEPC. A 2D cluster-based permutation t-test (Maris and Oostenveld 2007) with 3,000 permutations, a peak t threshold of p < 0.01, and a cluster threshold of p < 0.05 was used to identify significant increases in IEPC in the MEG data within this time window and frequency range, compared to the pre-event baseline. Baseline IEPC was calculated as the average IEPC between −400 and −100 ms relative to event onset in each frequency band.

In the four competing speech conditions, IEPC was calculated separately for target stream and competitors, using acoustic edges extracted from the audio data before mixing. IEPC of the two competitors for each listening condition was averaged.

To quantify the effects of speech cues and their interactions on speech tracking, we employed a modeling approach that required reduced data dimensionality. Our previous research (Oganian et al. 2023) used data simulation with an evoked response model to demonstrate that transient IEPC increases in the theta-band (4-8 Hz) constitute the primary driver of neural speech-tracking responses. Additionally, theta-band IEPC magnitude serves as a quantitative measure of neural responses. Based on these established findings, we focused on theta-band (4-8 Hz) IEPC measurements to investigate speech-tracking differences between groups and the effects of speech cues. The average IEPC in the theta band (4-8 Hz) was calculated by averaging IEPC values within this frequency band across the duration of a single cycle for each frequency. The topographic distribution of theta-band IEPC was visualized by plotting the average IEPC in the 4-8Hz range between 0 and 200 ms for each listening condition and group.

### 2.6. Caregiver questionnaire

The ECLiPS (Barry and Moore 2021; Denys et al. 2024) is a 38-item questionnaire in which caregivers rate their child’s behaviors on a five-point scale from strongly disagree to strongly agree. Individual item ratings are first age-scaled before being averaged to derive scores on five subscales (speech and auditory processing, environmental and auditory sensitivity, language/literacy/laterality, memory and attention, and pragmatic and social skills) with 6-9 items each. The subscales are aggregated into Language, Listening, Social, and Total composite scores, all standardized with a population mean of 10 (SD = 3)(Barry et al. 2015).

### 2.7. Cognition (NIH Toolbox)

Cognitive abilities were evaluated using the NIH Toolbox for the Assessment of Neurological and Behavioral Function, Cognition Domain (Weintraub et al. 2013b). Testing was completed online or via an iPad app in a sound-attenuated booth or quiet room.

The cognitive battery encompassed multiple domains: selective attention (Flanker test), episodic memory (Picture Sequence Memory Test), working memory (List Sorting Working Memory Test), executive function (Dimensional Change Card Sort Test), processing speed (Pattern Comparison Processing Speed Test), and verbal ability (Picture Vocabulary and Reading Recognition Tests). Three composite scores were derived from these assessments: fluid cognition, crystallized cognition, and overall cognition. For participants with missing behavioral data (4% of cases), values were imputed using data from their previous visits to the parent study. No imputation was performed for imaging data.

### 2.8. Statistical analysis

Statistical analyses were performed using R (version 4.3.0; R Core Team, 2023) in RStudio (version 2023.06.0 Build 421) and Matlab (version R2023b; The MathWorks Inc., Natick, MA, 2023). Demographic variables, including age, sex, race, ethnicity, and maternal education, were compared between the TD and LiD groups using independent samples t-tests for continuous variables and chi-square or Fisher’s exact tests for categorical variables. Statistical significance was set at *p* < 0.05.

Linear mixed-effects models were employed to examine the effects of group (TD vs. LiD), speech context (clear vs. competing speech), the presence of talker and spatial cues, and their interactions on both behavioral and neural outcomes. For behavioral data, the percentage of correct answers on comprehension questions served as the dependent variable. For neural data, theta IEPC to target speech and competitors were the primary outcomes. Separate models were constructed to assess the effects of speech context and cue type, with participant included as a random effect and age as a covariate. Additionally, to evaluate the influence of cognitive ability on comprehension performance during competing speech listening, a mixed-effects model was fitted with percent-correct comprehension as the repeated-measures outcome, NIH Toolbox Total Composite score as a fixed effect predictor, group, competing speech condition, and age as fixed covariates, and subject as a random intercept.

To examine the association between neural tracking and listening skills, linear regression analyses were performed with the ECLiPS total scaled score as the dependent variable and theta IEPC as the independent variable. Initial simple regression analysis was conducted for the competing speech condition incorporating both talker and spatial cues, as it represents the most ecologically valid real-world listening scenario (Auerbach and Gritton 2022; Cameron and Dillon 2007). Subsequently, participant group (LiD vs. TD) and recording age were included as covariates to control for potential bias stemming from inherent differences in ECLiPS scores due to initial group classification. Model performance was assessed using adjusted R² values. Additionally, the unique contribution of theta IEPC to explaining variance in ECLiPS scores was quantified by calculating partial R², which measures the incremental variance explained by theta IEPC beyond the variance accounted for by age and group alone. Exploratory analyses were conducted using similar regression approaches to identify associations between theta IEPC and NIH Toolbox composite scores, subscores, and individual ECLiPS subdomains.

## 3. RESULTS

### 3.1. Differences in questionnaire scores and behavioral performance between LiD and TD groups

As anticipated, the ECLiPS total scaled score and all subscores were lower in the LiD group compared to the TD group (Table 1, Supplemental Table S4). Overall cognitive performance, assessed by the NIH Toolbox Total Composite Score (Weintraub et al. 2013a; Weintraub et al. 2013b), and most subscores were also lower in the LiD group (Table 1, Supplemental Table), consistent with previous findings when the study group was younger (Petley et al. 2021; Kojima et al. 2023).

**Table 1.** Participant numbers and demographic information.

|  | TD (n = 25) | LiD (n = 21) | <sup>3</sup><br>*p* value <sup>4</sup> |
| --- | --- | --- | --- |
| Age, mean (SD) | 15.1 (2.3) | 15.4 (2.4) | .61 |
| Female (%) | 10 (44) | 11 (52) | .77 |
| Race (%) |  |  | .61 |
| Black | 7 (28) | 4 (19) |  |
| White | 17 (68) | 15 (71) |  |
| Mixed | 1 (4) | 2 (10) |  |
| Hispanic (%) | 0 (0) | 1 (5) | .46 |
| Maternal education (%) |  |  | .35 |
| High school | 2 (8) | 3 (14) |  |
| Some college | 2 (8) | 4 (19) |  |
| College | 17 (68) | 9 (43) |  |
| Postgraduate | 4 (16) | 5 (24) |  |
| ECLiPS Total Scaled Score, mean (SD) | 9.8 (2.6) | 4.0 (2.7) | <.0001 |
| NIH-Toolbox Total Composite, mean (SD) | 115 (20) | 96 (20) | .003 |

During the competing speech listening task, the percentage of correct responses to comprehension questions was lower in the LiD group compared to the TD group (β = −11.4, SE = 4.5, t = −2.6, p = .014; Figure 2). This group difference was consistent across conditions, with no interaction effects between group and either talker or spatial cues observed. Both groups demonstrated reduced comprehension accuracy under competing speech relative to the clear speech (β = −6.3, SE = 2.1, t = −3.0, p = .003).

**Figure 2.**
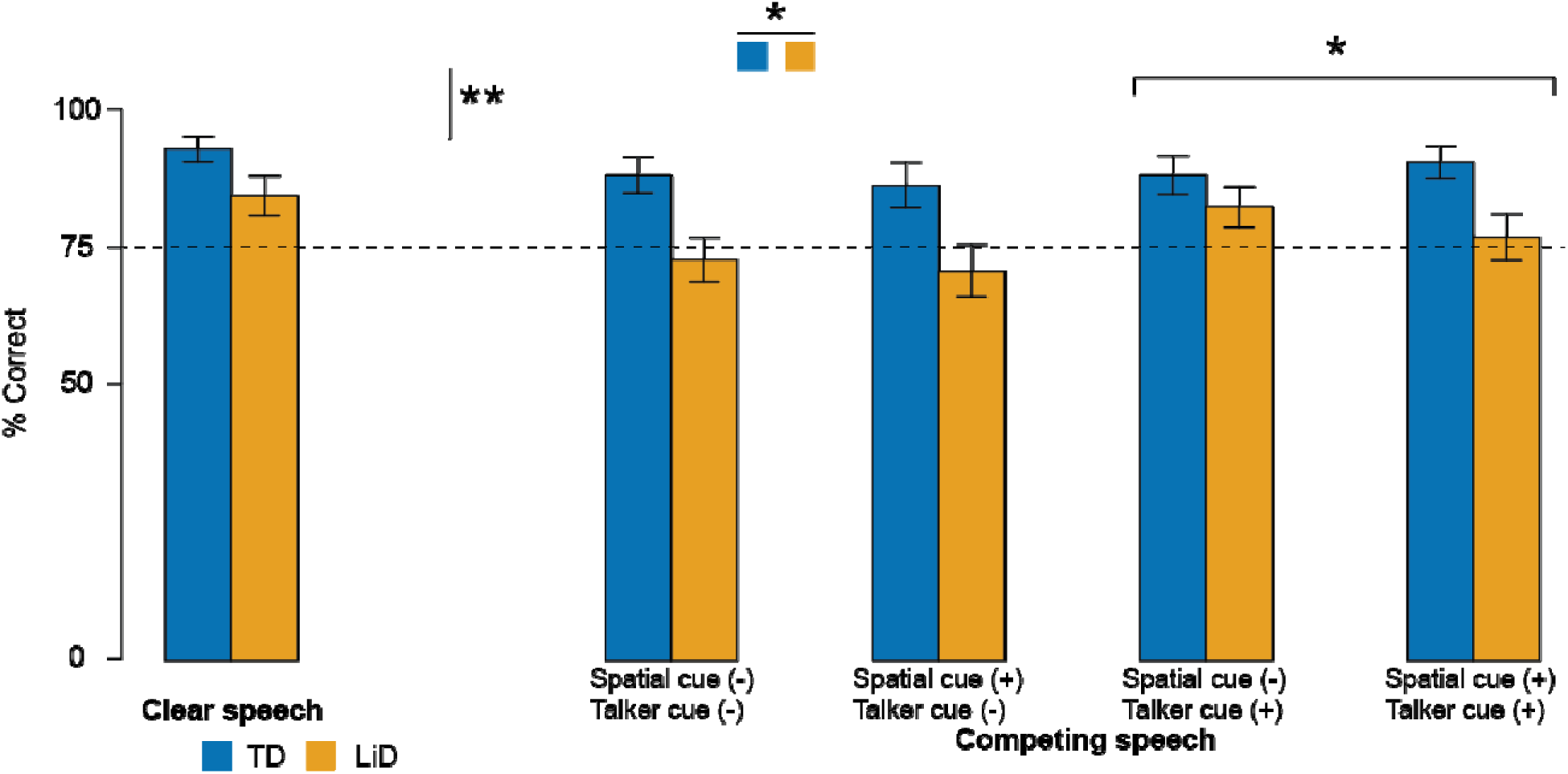
Comprehension accuracy across listening conditions in adolescents with listening difficulty (LiD) and typically developing (TD) controls. Participants answered multiple-choice questions about target speech presented either in isolation (clear speech) or with competing speech streams. Competing speech conditions varied systematically by the presence (+) or absence (-) of talker cues (different vs. same voice) and spatial cues (different vs. same location). Statistical analysis revealed significantly lower comprehension in the LiD group compared to TD controls, with both groups showing reduced performance during competing speech conditions. The presence of talker cues improved comprehension accuracy across groups. Blue and orange bars represent TD and LiD participants, respectively. Error bars show ± one standard deviation. Asterisks indicate significant differences (*p < .05, **p < .01). The dashed horizontal line at 75% accuracy is included as a visual aid and does not represent a statistical threshold or reference value.

Additionally, comprehension significantly improved when talker cues were present compared to absent (β = 4.6, SE = 1.8, t = 2.5, p = .014), suggesting talker cues facilitated comprehension in challenging listening scenarios. Finally, the percent correct responses to comprehension questions were consistently and significantly associated with NIH Toolbox Total Composite scores (β = 0.47, SE = 0.1, t = 4.7, p < .001) across both groups and all listening conditions.

### 3.2. Reduced neural speech tracking in LiD during competing speech listening

Average evoked responses to sentence onset, quantified as the M100 response, did not differ between TD and LiD participants (cluster-based permutation testing > .05; Figure 1C), indicating comparable basic auditory processing across participants. NST, measured as theta-band (4 - 8Hz) IEPC, showed significant increases relative to pre-event baseline during clear speech listening in both TD and LiD groups (cluster-based permutation testing, p < .05; Figure 3B, Supplemental Figure S2). However, group differences emerged under competing speech conditions. While the TD group consistently demonstrated elevated theta-band IEPC across all competing speech conditions (cluster-based permutation testing, p < .05), the LiD group exhibited no significant IEPC enhancement during these challenging listening conditions.

**Figure 3.**
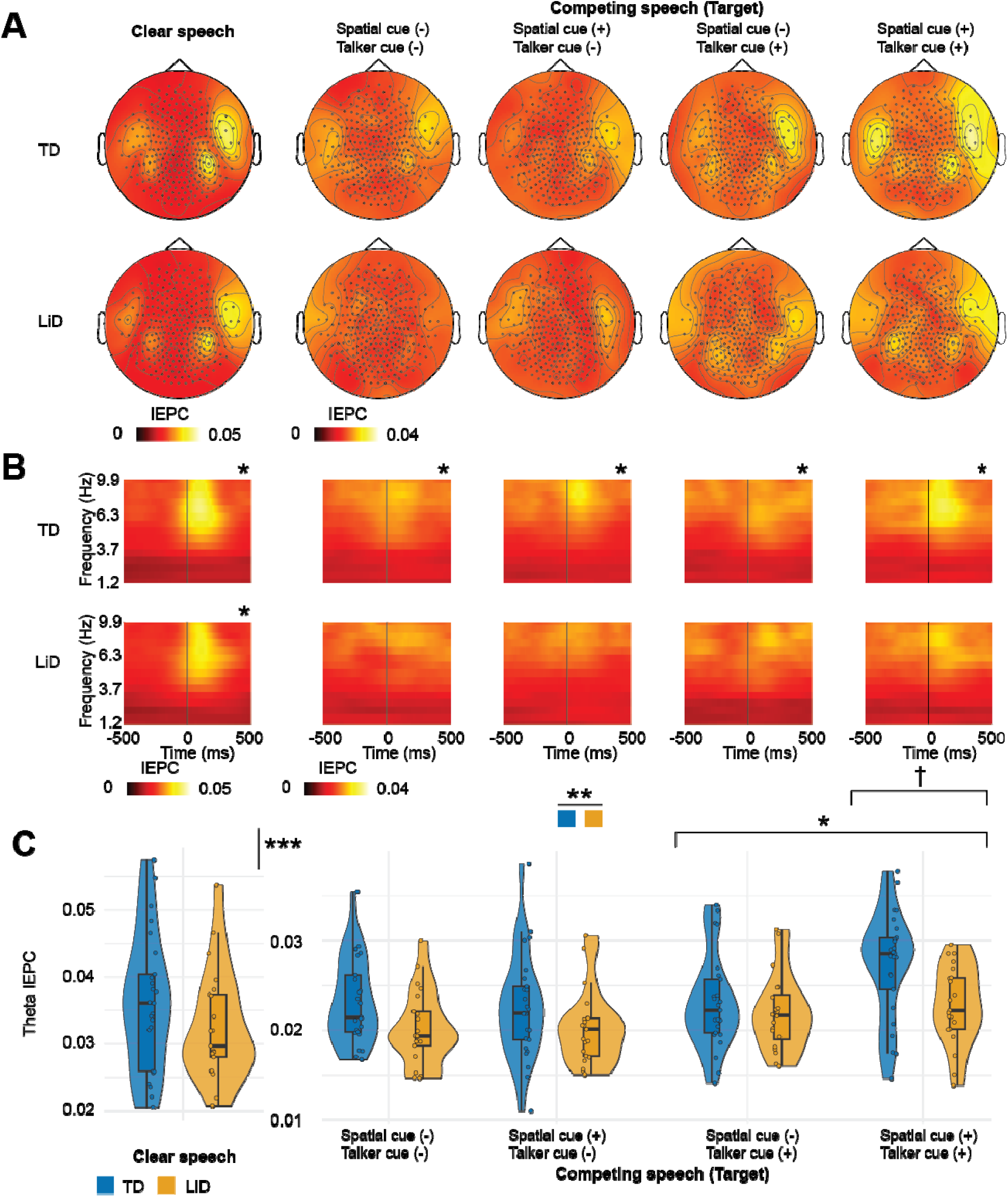
Neural speech tracking of target speech during clear and competing speech conditions. **A.** Topographic maps showing theta-band IEPC to acoustic edges in target speech for TD and LiD groups. Maps are shown for clear speech (leftmost) and four competing speech conditions with varying spatial and talker cues (right). **B.** Time-frequency representations of IEPC patterns relative to acoustic edges (time 0) for each condition. The asterisk (*) at the top indicates a statistically significant cluster compared to baseline (p < .05, cluster-based permutation testing). **C.** Violin plots showing average theta-band IEPC across conditions. Neural speech tracking was significantly reduced in competing speech compared to clear speech conditions, and the LiD group showed consistently lower IEPC than TD controls. Talker cues enhanced neural speech tracking, whereas spatial cues alone did not. However, the combination of talker and spatial cues synergistically increased theta IEPC. Asterisks indicate significant differences (*p < .05, **p < .01, ***p < .001). The dagger (†) indicates a significant interaction between spatial and talker cues.

Despite this difference between groups, direct statistical comparisons in the 2D heatmaps between LiD and TD participants did not reach significance across listening conditions. These findings suggest reduced robustness of NST in children with LiD under adverse listening conditions.

### 3.3. Reduced theta phase-locking to target speech in LiD and during competing speech, with enhancement by combined spatial and talker cues

Analysis of average theta-band (4-8 Hz) IEPC revealed reduced NST to the *target* speech during competing speech compared to clear speech conditions (Figure 3C; β = −1.2 × 10^2^, SE = 8.9 × 10, t = −13, p < .001). This indicates substantial interference from competing auditory streams on the neural processing of target speech. However, the magnitude of this disruption did not differ significantly between the TD and LiD groups, as evidenced by the non-significant interaction between participant group and auditory competition (β = 6.5 × 10, SE = 1.8 × 10^3^, t = 0.4, p = .715). The LiD group consistently exhibited lower theta IEPC compared to the TD group across both clear and competing speech conditions (β = −3.1 × 10^3^, SE = 1.1 × 10^3^, t = −2.8, p = .008). Collectively, these findings suggest a generalized deficit in neural synchronization to target speech among individuals with LiD, irrespective of the presence of auditory competition.

In conditions with competing speech, the presence of talker cues enhanced theta IEPC (β = 2.2 x 10^-3^, SE = 6.7 x 10^-4^, t = 3.4, p = .001; Figure 3C). A similar, albeit non-significant, trend was observed for spatial cues (β = 1.2 x 10^-3^, SE = 6.7 x 10^-4^, t = 1.8, p = .08). Additionally, there was a significant interaction between talker and spatial cues (β = 3.2 x 10^-3^, SE = 1.3 x 10^-3^, t = 2.4, p = .018). Post hoc pairwise comparisons clarified this interaction, demonstrating that neither spatial cues alone (p = .974) nor talker cues alone (p = .896) significantly enhanced theta IEPC relative to the baseline condition (no talker or spatial cues). In contrast, the simultaneous combination of both cue types yielded significant enhancements compared to baseline (p = .002), as well as compared to spatial cues alone (p < .001) and talker cues alone (p = .019). These results highlight a synergistic relationship between spatial and talker cues, where significant neural enhancement occurs predominantly when both cues are concurrently available. Thus, the previously reported main effect of talker cues appears predominantly driven by this synergistic interaction.

While the formal three-way interaction (talker cue × spatial cue × group) did not reach significance (β = −4.0 × 10^3^, SE = 2.6 × 10^3^, t = −1.5, p = .130), likely reflecting limited statistical power, post hoc analyses revealed notable group-specific differences. Specifically, the synergistic interaction between talker and spatial cues significantly enhanced theta IEPC in the TD group (β = 5.0 × 10^3^, SE = 2.0 × 10^3^, t = 2.5, p = .016), consistent with the main findings described above. In contrast, this interaction effect was not statistically significant in the LiD group (β = 9.7 × 10, SE = 1.6 × 10^3^, t = 0.6, p = .544). These results suggest a robust combined benefit of spatial and talker cues among TD individuals, while the effectiveness of cue combination appears attenuated or absent in LiD participants, although this interpretation remains tentative given the lack of significant three-way interaction in this sample. Additionally, no significant correlation was observed between theta IEPC and the accuracy of comprehension responses.

In summary, our analysis of theta IEPC related to *target* speech processing yielded three key findings: (1) adolescents with LiD exhibited broadly reduced NST compared to their TD peers, irrespective of auditory competition; (2) NST is significantly impaired during competing speech conditions relative to clear speech in both groups; and (3) simultaneous provision of spatial and talker cues synergistically enhanced NST, an effect that appears more robust in the TD group.

### 3.4. Comparable neural speech tracking of competitors in TD and LiD groups supports selective deficits of target speech processing in LiD

To further elucidate the mechanisms underlying LiD, we examined neural tracking of *competitor* speech streams during competing speech conditions (Figure 4). In the TD group, theta-band IEPC increased relative to the pre-event baseline across all competing speech conditions (Figure 4B, top row; cluster-based permutation testing, p < .05; Supplemental Figure S2), indicating robust neural tracking of competitor speech. Similarly, the LiD group demonstrated elevated theta-band IEPC predominantly under conditions involving spatial cues, suggesting that spatial separation may facilitate competitor tracking.

**Figure 4.**
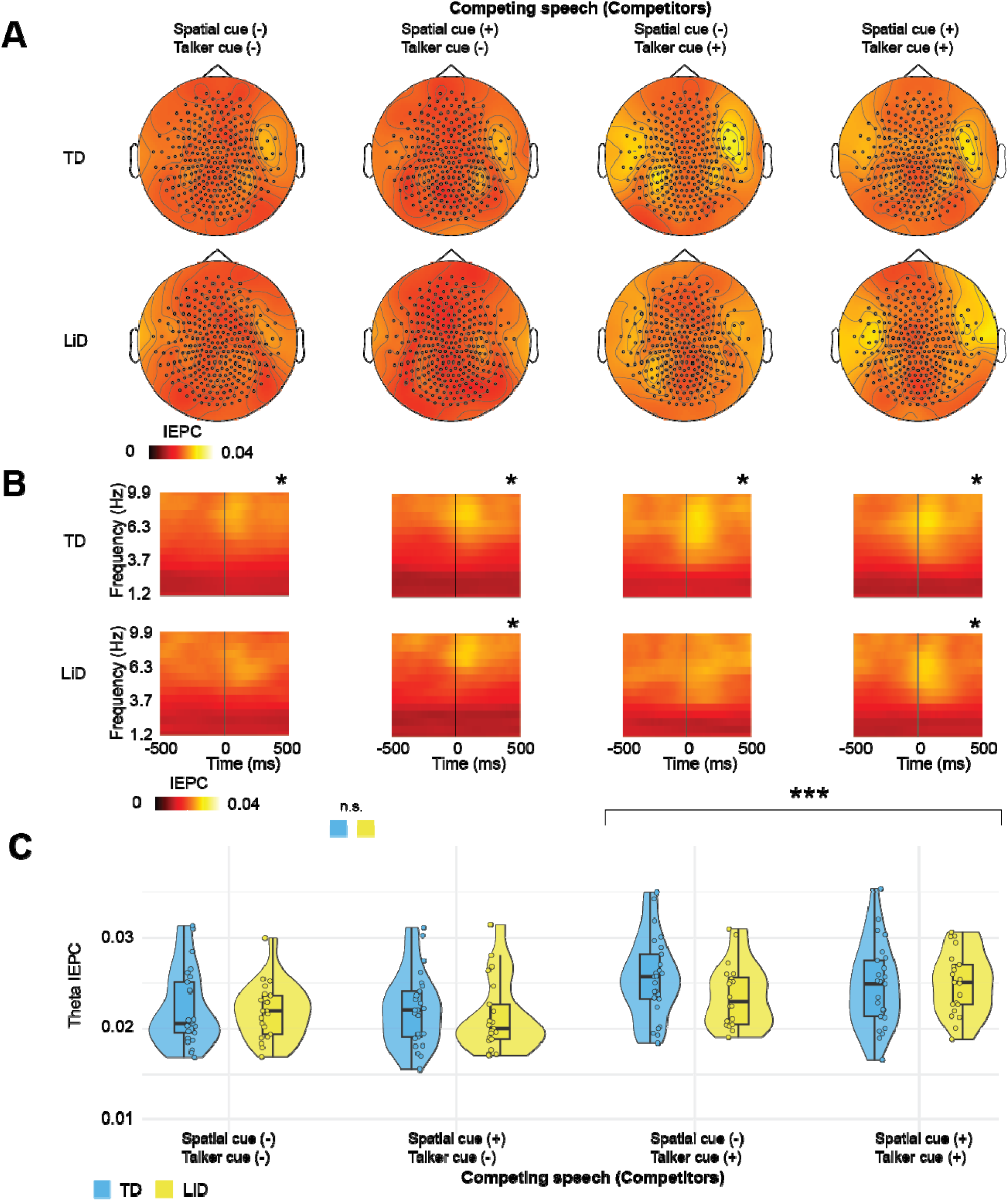
Neural speech tracking of competitors in TD adolescents and those with LiD. **A.** Topographic distributions of theta-band IEPC following acoustic edges in competitors across four competing speech conditions, varying in spatial and talker cues. **B.** Time-frequency IEPC patterns for each condition relative to the acoustic edge (time 0). The asterisk (*) at the top indicates a statistically significant cluster compared to baseline (p < .05, cluster-based permutation testing). **C.** Distribution of theta-band IEPC values for TD (light blue) and LiD (yellow) groups across conditions. Statistical analysis revealed no differences between groups in competitor tracking, contrasting with the reduced target speech tracking observed in LiD. Both groups showed enhanced tracking when talker cues were present. Significant effects across conditions are indicated by *** (p < .001).

Consistent with our hypothesis, theta IEPC to competitor speech did not differ between TD and LiD groups (β = −7.9 x 10^-4^, SE = 7.8 x 10^-4^, t = −1.0, p = .311) (Figure 4C). A combined analysis of theta IEPC for both target and competitor speech revealed a significant interaction between participant group and speech stream type (target vs. competitor; β = −2.1 x 10^-3^, SE = 8.4 x 10^-4^, t = −2.5, p = .012). This interaction highlights that group differences in neural speech tracking are specifically modulated by whether the auditory input serves as target or competitor.

As observed with target speech, theta IEPC to competitor speech increased with talker cue availability (β = 2.9 x 10^-3^, SE = 4.8 x 10^-4^, t = 6.0, p < .001), and this effect did not differ by participant group. Unlike responses to target speech, no interaction emerged between spatial and talker cues in neural tracking of competitor speech (β = 4.9 x 10^-4^, SE = 9.6 x 10^-4^, t = 0.5, p = .610). Additionally, no significant difference was observed between theta IEPC to target versus competitor speech overall (β = 8.0 x 10^-4^, SE = 9.2 x 10^-4^, t = 0.9, p = .389).

Collectively, these results indicate that neural tracking of competitor speech streams during competing listening conditions is comparable between adolescents with LiD and their TD peers. Together with the findings of reduced theta IEPC to target speech in the LiD group presented in Section 3.3, this pattern supports our hypothesis that LiD symptoms in our cohort primarily reflect selective impairments in processing target speech within complex auditory environments.

### 3.5. Neural speech tracking correlates with caregiver-reported listening difficulty

We hypothesized that neural tracking of target speech (theta IEPC) during competing speech conditions would correlate positively with caregiver-reported listening skills, as assessed by the ECLiPS total scaled score. Initial simple linear regression analysis for the ecologically valid condition incorporating both talker and spatial cues revealed a significant positive association between theta IEPC and ECLiPS scores (β = 326, SE = 92, t = 3.6, p < .001; Figure 5, dashed line). After controlling for participant group (LiD vs. TD) and recording age, this association remained significant (β = 164, SE = 76, t = 2.1, p = .037; Figure 5, solid lines), indicating higher theta IEPC values correspond to better listening skills across both groups.

**Figure 5.**
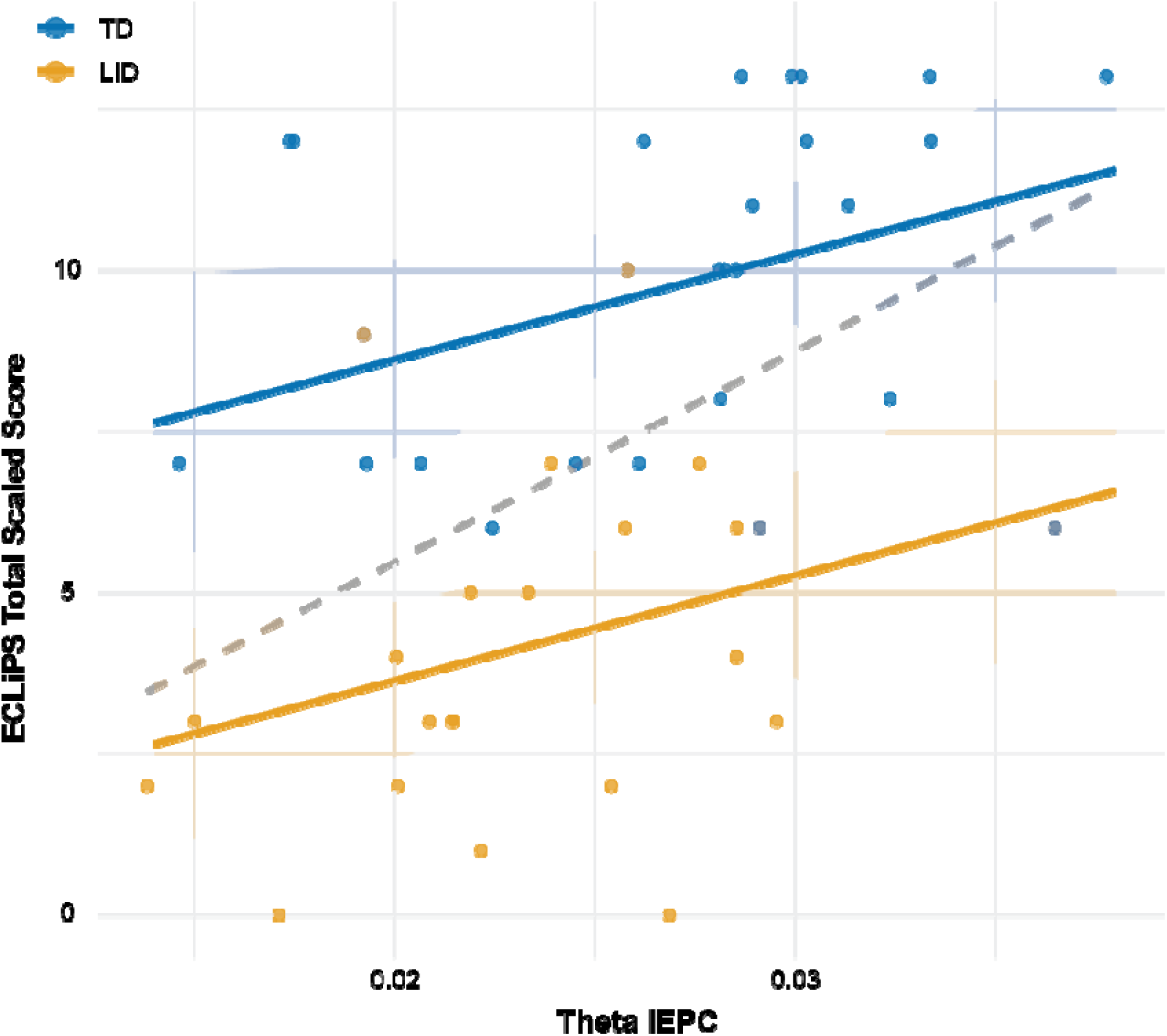
Relationship between neural speech tracking and caregiver-reported listening ability during a competing speech condition. The scatter plot illustrates the linear association between theta IEPC values and ECLiPS Total Scaled Scores for participants with LiD (orange) and TD (blue), recorded during the competing speech condition incorporating both talker and spatial cues. The gray dashed line indicates the overall regression line across groups without covariate adjustment. Solid regression lines with shaded areas (95% confidence intervals) depict age-adjusted linear relationships within each group separately. Higher theta IEPC values were significantly associated with better caregiver-reported listening abilities across both groups after adjusting for participant age and group (p = .037), while the LiD group consistently demonstrated lower ECLiPS scores compared to TD controls (p < .001).

The adjusted regression model explained 56.9% of the variance in ECLiPS scores, with theta IEPC uniquely accounting for an additional 10% of the variance (partial R^2^ = .10). No significant associations emerged in other competing speech conditions.

Exploratory analyses examining associations between theta IEPC and NIH Toolbox composite scores, subscores, and ECLiPS subdomains revealed a borderline significant relationship with NIH Toolbox List Sorting (overall model R^2^ = 0.165, model p = .054; theta IEPC p = .039). Significant associations between IEPC were identified with several ECLiPS subdomains, including Environment and Auditory Sensitivity (R^2^ = 0.476, model p < .001; theta IEPC p = .047), Pragmatic and Social Skills (R^2^ = 0.428, model p < .001; theta IEPC p = .037), Listening aggregate (R^2^ = 0.516, model p < .001; theta IEPC p = .028), and Social aggregate (R^2^ = 0.479, model p < .001; theta IEPC p = .017). No other NIH Toolbox composites or ECLiPS subscales demonstrated significant associations.

These results indicate that neural speech tracking, specifically theta IEPC during ecologically valid listening conditions, is significantly related to caregiver-reported listening ability. Furthermore, associations with specific ECLiPS subdomains suggest that neural speech tracking may reflect underlying auditory sensitivity and social-pragmatic skills.

## 4. DISCUSSION

In this study, we investigated the neural mechanisms underlying LiD by recording MEG during a cocktail party task in adolescents with LiD and their TD peers. Adolescents with LiD exhibited reduced NST of target speech compared to TD adolescents, irrespective of the presence of competing speech. Importantly, the strength of NST to target speech correlated positively with caregiver-reported listening abilities across both groups, indicating that theta-band phase-locking responses provide a meaningful neurophysiological correlate of real-world listening performance. Although NST to target speech was diminished in adolescents with LiD, their neural responses to competitor speech stream were comparable to those of TD peers, supporting the hypothesis that LiD reflects selective deficits in processing attended speech rather than enhanced tracking of competitor speech or generalized auditory disengagement. Furthermore, we observed that the simultaneous presence of talker and spatial cues synergistically enhanced NST to target speech, an effect more pronounced in the TD group. The absence of this synergistic effect in NST to competitor speech streams further highlights the specificity and importance of attentional mechanisms in utilizing these auditory cues to segregate and attend to target speech in complex listening environments.

### 4.1. Association of neural speech tracking with listening difficulty

This study provides the first empirical evidence linking LiD symptoms to deficits in NST. The correlation between NST of ecologically relevant, cue-rich target speech in a competing speech environment and caregiver-reported symptom scores is particularly notable, given the broad inclusion criteria based on caregiver ratings. LiD is a heterogeneous condition encompassing impairments in hearing, auditory processing, phoneme recognition, cognition, and language, involving both bottom-up sensory and top-down cognitive mechanisms (Dillon and Cameron 2021). Given this multifactorial nature, identifying an objective neurophysiological measure associated with subjective symptom severity marks a significant advance in understanding the neural bases of LiD.

Theta IEPC, as analyzed here, specifically indexes auditory cortical phase-locking to rapid amplitude transitions (acoustic edges) in the speech envelope. Precise phase-locking to acoustic edges is crucial for parsing syllabic boundaries and prosodic stress, enabling listeners to segregate target speech from competing auditory streams (Ding and Simon 2014; Doelling et al. 2014). This mechanism aligns closely with caregiver assessments of listening ability, particularly the Environment & Auditory Sensitivity subscale and the broader Listening Aggregate of the ECLiPS. Adolescents who demonstrate stronger neural edge-locking are perceived by caregivers as better able to manage daily acoustic environments. Consistent associations between neural envelope tracking and behavioral listening outcomes have previously been reported in other populations, reinforcing our findings (Vanthornhout et al. 2018; Fuglsang et al. 2020).

Although the observed association between theta IEPC and pragmatic/social skills might initially appear speculative, given the complex cognitive nature of those skills. However, pragmatic aspects of conversation heavily rely on accurate, real-time decoding of prosody, turn-taking cues, and speaker identification—processes partially mediated by amplitude edges captured by theta IEPC (Rimmele et al. 2018). Poor encoding of these edges could therefore reduce sensitivity to prosodic cues critical for interpreting emotional tone, irony, or conversational intent, potentially making complex multi-talker interactions challenging and socially stressful. Over time, these challenges may lead to social withdrawal behaviors, influencing caregivers’ perceptions of pragmatic competence negatively (Sharma et al. 2009; Moore et al. 2010). Indeed, auditory processing deficits have previously been associated with pragmatic and social difficulties, including traits within the autism spectrum, supporting a hypothesized sensory-to-social cascade (Dawes and Bishop 2009). While these secondary analyses provide mechanistically plausible pathways connecting auditory edge detection and social-pragmatic outcomes, they are exploratory. Future studies explicitly designed with pre-registered hypotheses will be necessary to confirm these preliminary insights definitively.

### 4.2. Neural mechanisms of competing speech processing in children with LiD

Adolescents with LiD demonstrated normal NST to unattended competitor speech streams; the significant group difference was attributed primarily to their diminished ability to enhance NST to the attended target stream. This selective deficit aligns with the view that LiD predominantly reflects impairments in selective auditory gain control mechanisms rather than difficulties in inhibiting distracting stimuli or generalized speech processing challenges. In typical listeners, top-down attention dynamically amplifies cortical tracking of target speech in the theta frequency range, while representations of competing background streams remain relatively unaffected (Kerlin et al. 2010; Zion Golumbic et al. 2013). Our findings suggest that this attentional amplification is compromised in adolescents with LiD, despite intact baseline encoding of the competing speech envelope. Such deficits imply inadequate modulatory support from higher-order cognitive control networks to the auditory cortex (Shinn-Cunningham and Best 2008; Fritz et al. 2007).

This deficit was most evident when both talker-identity and spatial-location cues were concurrently available—conditions under which TD adolescents exhibited a synergistic enhancement of NST to target speech. Efficient utilization of multiple auditory cues requires rapid integration of spectro-temporal voice characteristics and spatial localization, processes reliant on coordinated activity within a distributed fronto-parietal attentional network that biases sensory cortices (Hill and Miller 2010; Michalka et al. 2016). The observed deficiency in cue integration among adolescents with LiD, therefore, highlights possible inefficiencies in multimodal feature binding rather than purely sensory encoding limitations. Such integration deficits may originate from developmental immaturity or disrupted long-range oscillatory coupling between auditory cortical regions and control hubs such as the inferior parietal lobule and dorsolateral prefrontal cortex, circuits known to mature through adolescence and influenced by auditory experience (Chai et al. 2016; Power et al. 2010; Heinrichs-Graham et al. 2022).

Results presented here reconcile previous inconsistent findings where children with LiD exhibited normal auditory brainstem or early cortical responses to simple acoustic stimuli but encountered significant challenges in complex auditory environments (Hunter et al. 2023). Consequently, our data advocate shifting clinical diagnostic assessments and therapeutic interventions toward enhancing selective attention mechanisms and cue integration skills rather than merely suppressing background noise. Specifically, interventions focusing on auditory cognitive training that progressively emphasizes the integration of spatial and talker cues, or neuromodulatory strategies targeting fronto-parietal theta-band synchrony, may yield improvements (Polley et al. 2006; Albouy et al. 2017). Future longitudinal studies should investigate whether enhancing attentional gain and cue integration during critical developmental windows can reduce subsequent academic and social impairments associated with LiD.

### 4.3. Methodological considerations in edge-based neural speech tracking analysis

An edge-centric approach to calculating NST confers several advantages. First, by isolating phase-locking specifically to rapid amplitude transitions—acoustic edges—it grounds our analysis in an early, modality-specific auditory processing stage conserved across both developmental stages and diverse clinical populations (Doelling et al. 2014; Gross et al. 2013; Van Hirtum et al. 2023; Vander Ghinst et al. 2019). Anchoring analyses at this sensory coding level reduces confounds stemming from higher-order linguistic or cognitive factors, thus attributing observed group differences directly to fundamental sensory mechanisms. Second, the analytic pipeline is intentionally streamlined: the metric relies primarily on band-limited filtering and circular statistics, circumventing the need for time-aligned linguistic annotations or complex multivariate fitting procedures. This methodological parsimony enhances reproducibility and facilitates validation across research sites (Kösem and van Wassenhove 2016), laying the groundwork for eventual clinical translation and assessment standardization.

Focusing on acoustic edges also enhances the interpretability and theoretical clarity of our findings. Since the theta-band response primarily reflects acoustic onsets, diminished NST to target speech can be succinctly attributed to impaired selective amplification processes, rather than complex phonemic, lexical, or semantic mismatches (Ding and Simon 2014; Doelling et al. 2014). Such sensory-level specificity complements, rather than competes with, more sophisticated multivariate temporal response function (mTRF) approaches that incorporate higher-order linguistic features (Crosse et al. 2016). Establishing a robust foundational correlate of selective attention at the sensory processing level provides a necessary baseline from which subsequent linguistic and cognitive effects can be systematically investigated, thereby ensuring that later-stage analyses are not confounded by uncharacterized sensory deficits.

However, our reliance on edge-based metrics has limitations. Future research should expand beyond this specific sensory measure to incorporate complementary approaches. For instance, mTRF analyses that independently parameterize phonemic, lexical, and semantic speech features would further elucidate how downstream language networks build upon or compensate for the sensory deficits identified here (Di Liberto et al. 2015; Broderick et al. 2018; Bolt and Giroud 2024). Such analyses could potentially uncover distinct neural signatures associated specifically with comprehension accuracy, executive functioning, or broader cognitive abilities—domains that were intentionally not explored in depth within our current analysis. To address this gap, we are currently conducting parallel mTRF studies within the same participant cohort, linking higher-order neural speech-tracking components to NIH Toolbox cognitive scores and behavioral comprehension performance. These ongoing investigations aim to determine whether targeted auditory-attention training can remediate deficits in both edge-locked sensory encoding and higher-order linguistic processing.

### 4.4 Limitations

While this study provides novel insights into the neural mechanisms underlying LiD, several methodological limitations should be acknowledged. First, the use of a fixed SNR of 5 dB between target and competing speech streams, though informed by prior studies (Vander Ghinst et al. 2021; Hausfeld et al. 2021) and preliminary testing, may have constrained our ability to fully characterize the effects of spatial and talker cues or potential differences in cue utilization between TD adolescents and those with LiD. Additionally, as this was an exploratory study, our sample size may have been insufficient to detect subtle group differences. Nevertheless, we successfully identified group differences between LiD and TD participants and linear association between theta IEPC and ECLiPS scores during competing speech listening. Future studies would benefit from examining multiple SNR levels and recruiting larger sample sizes to further elucidate these relationships.

### 4.5 Conclusion

This study provides new insights into the neural mechanisms underlying LiD, demonstrating that adolescents with LiD exhibit selective deficits in neural tracking of attended speech, despite normal processing of competitor speech streams. Our findings indicate that LiD primarily arises from selective impairments in processing attended speech rather than heightened processing of competing sounds or general auditory disengagement. Correlation between neural speech tracking and caregiver-reported listening difficulties further validates this neurophysiological measure as a meaningful index of real-world listening challenges. Reduced ability of adolescents with LiD to integrate talker and spatial cues, compared to their TD peers, suggests that inefficiencies in cue integration may contribute to listening difficulties. Overall, this study advances our understanding of the neurobiological foundations of LiD and emphasizes the role of selective auditory attention in managing complex listening environments.

## Data Availability

All data produced in the present study are available upon reasonable request to the authors

## ACKNOWLEDGEMENTS

We extend our gratitude to KidNuz for generously allowing us to use their children’s news podcast as speech stimuli in this research project. This research was supported by grant 5R01DC014078 from the National Institute of Deafness and other Communication Disorders (DRM), 2UL1TR001425-05A1 from the National Center for Advancing Translational Sciences of the National Institutes of Health (KK), R21DC022030 from the National Institute on Deafness and Other Communication Disorders (KK), and by the Cincinnati Children’s Research Foundation. DRM received support from NIHR Manchester Biomedical Research Centre (NIHR203308).

---

**Supplemental Table S1.** Stimulus information.

| **Listening condition** | **Audio** | **Talker** | **Duration (sec)** | **Acoustic edge number** | **Acoustic edge frequency (Hz)** | **Fundamental frequency (Hz)** |
| --- | --- | --- | --- | --- | --- | --- |
| Clear | Target stream | Tori | 332 | 1675 | 6.6(2.9) | 204(69) |
| Spatial cue (-),<br>Talker cue (-) | Target stream | Tori | 355 | 1826 | 6.9(3.1) | 201(69) |
|  | Competitor 1 | Tori |  | 1770 | 6.7(2.9) | 199(68) |
|  | Competitor 2 | Tori |  | 1826 | 6.7(2.9) | 198(68) |
| Spatial cue (+),<br>Talker cue (-) | Target stream | Tori | 355 | 1874 | 6.9(3.0) | 201(69) |
|  | Competitor 1 | Tori |  | 1829 | 6.9(3.0) | 194(65) |
|  | Competitor 2 | Tori |  | 1926 | 7.1(3.0) | 197(70) |
| Spatial cue (-),<br>Talker cue (+) | Target stream | Tori | 345 | 1767 | 6.7(2.9) | 199(67) |
|  | Competitor 1 | Kim |  | 1615 | 6.4(3.0) | 207(71) |
|  | Competitor 2 | Kim |  | 1640 | 6.4(2.9) | 205(71) |
| Spatial cue (+),<br>Talker cue (+) | Target stream | Tori | 356 | 1802 | 6.7(3.0) | 200(69) |
|  | Competitor 1 | Kim |  | 1744 | 6.7(3.0) | 203(67) |
|  | Competitor 2 | Kim |  | 1681 | 6.5(3.0) | 203(70) |

\*All values are mean (SD) unless otherwise noted.

**Supplemental Table S4.** Questionnaire and Cognitive testing scores.

| Testing measures<sup>*</sup> | TD (n = 25) | LiD (n = 21) | p value |
| --- | --- | --- | --- |
| ECLiPS scores |  |  |  |
| Speech and auditory processing | 10.2 (3.0) | 4.1 (3.2) | < .001 |
| Environmental and auditory sensitivity | 10.4 (2.9) | 5.7 (3.6) | <.001 |
| Language/literacy/laterality | 10.0 (2.3) | 4.3 (2.8) | <.001 |
| Memory and attention | 7.7 (1.7) | 5.2 (2.2) | <.001 |
| Pragmatic and social skills | 11.0 (3.4) | 6.3 (2.8) | <.001 |
| Language | 8.2 (1.8) | 4.1 (2.3) | <.001 |
| Listening | 9.3 (2.6) | 4.7 (2.6) | <.001 |
| Social | 10.8 (3.2) | 5.7 (3.5) | <.001 |
| NIH Cognition Toolbox Scores |  |  |  |
| Picture vocabulary | 105 (16) | 94 (10) | .006 |
| Flanker test | 106(17) | 96 (20) | .08 |
| List sorting working memory | 110 (20) | 101 (19) | .13 |
| Dimensional change card sort test | 112 (22) | 97 (31) | .03 |
| Pattern comparison processing speed test | 116 (22) | 97 (31) | .03 |
| Picture sequence memory test | 110 (18) | 103 (16) | .16 |
| Oral reading recognition test | 110 (14) | 96 (15) | .003 |
| Fluid cognition composite | 117 (23) | 98 (26) | .01 |
| Crystallized cognition composite | 106 (23) | 95 (13) | .04 |

<sup>\*</sup>All values are mean (SD) unless otherwise noted.

**Supplemental Figure S1.**
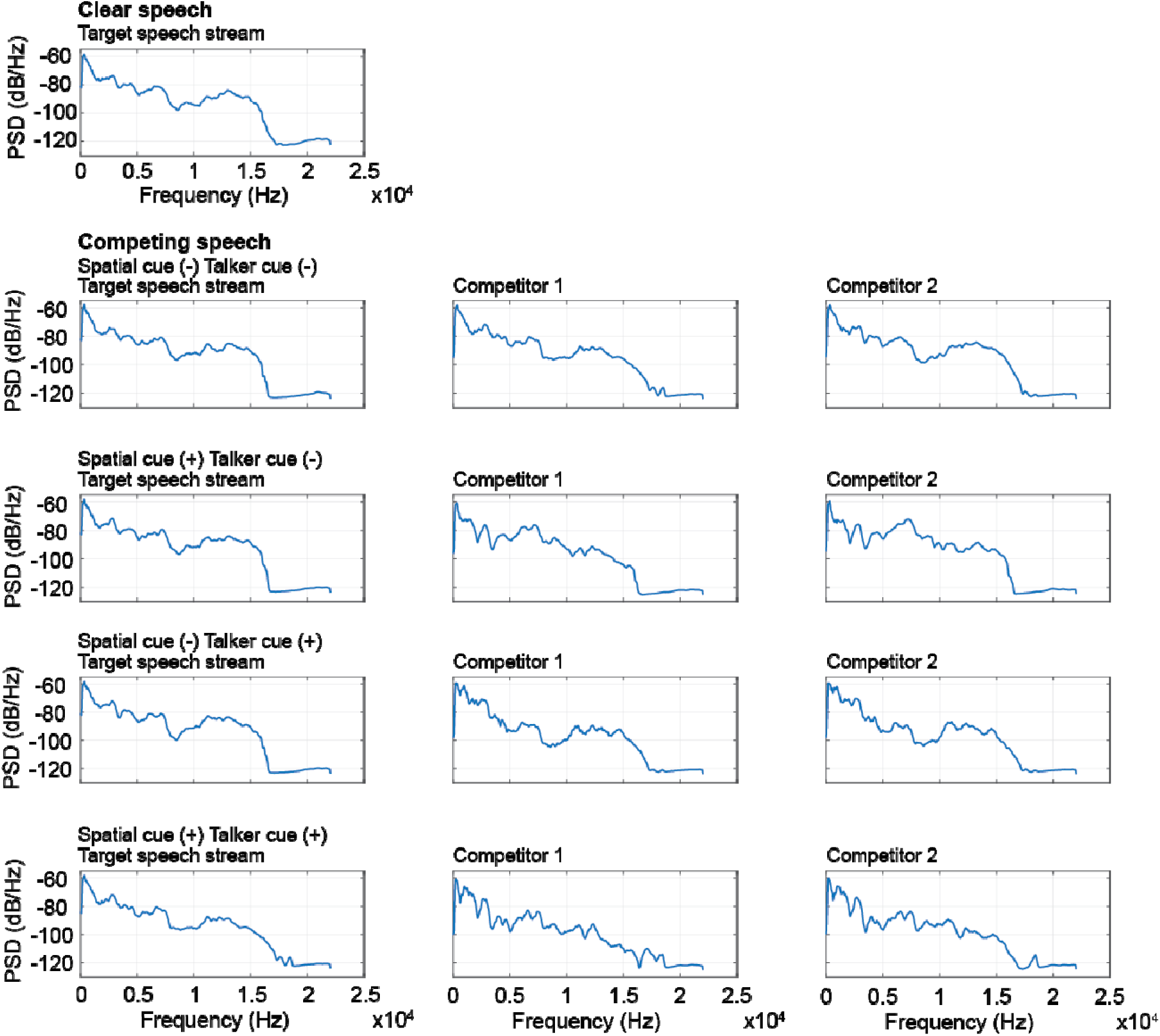
Power Spectral Density of Target Speech Stream and Competitors Across Experimental Conditions Power spectral density (PSD) measurements (dB/Hz) of auditory stimuli across frequencies (0-25 kHz). Top panel shows the target speech stream in clear speech condition. Lower panels display PSDs for the four competing speech conditions with varying spatial and talker cues: (1) Spatial cue (-) Talker cue (-), (2) Spatial cue (+) Talker cue (-), (3) Spatial cue (-) Talker cue (+), and (4) Spatial cue (+) Talker cue (+). For each competing condition, PSDs are shown for the target speech stream (left column) and two competitors (middle and right columns). All stimuli demonstrate comparable spectral characteristics with primary energy concentrated below 8 kHz and similar frequency roll-off patterns above 15 kHz, ensuring acoustic consistency across experimental conditions while manipulating spatial and talker identity cues.

**Figure S2.**
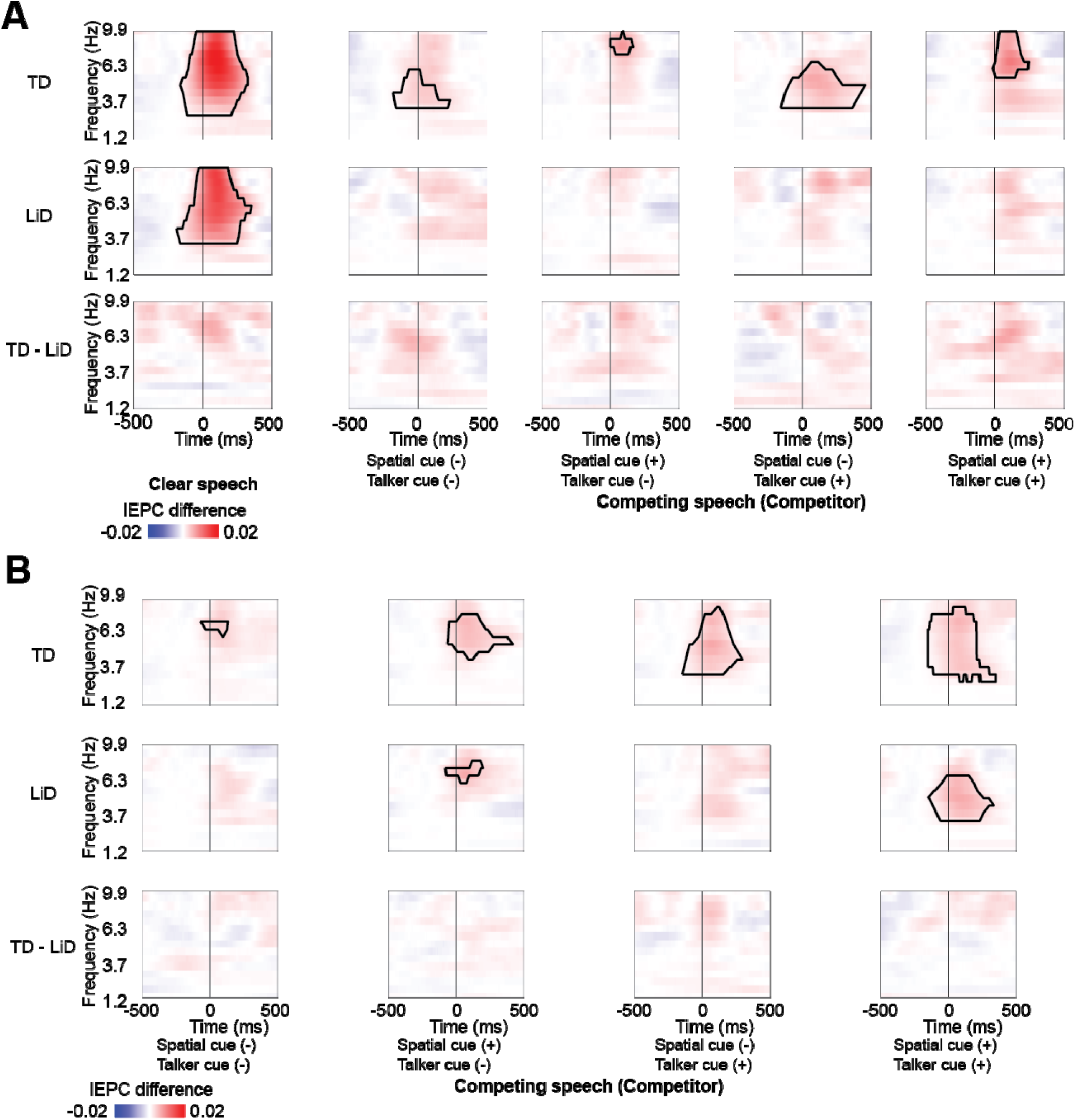
Group differences in theta-band IEPC to target speech and competitors. **A.** Time-frequency representations showing IEPC differences of the target speech stream during clear and four competing speech conditions. The top row illustrates IEPC increases from baseline in TD participants, the middle row represents LiD participants, and the bottom row displays TD– LiD subtraction maps. Contour lines indicate statistically significant clusters determined by cluster-based permutation testing (*p < .05). **B.** Time-frequency representations showing IEPC differences of competitors during four competing speech conditions. The top row illustrates IEPC increases from baseline in TD participants, the middle row represents LiD participants, and the bottom row displays TD–LiD subtraction maps. Contour lines indicate statistically significant clusters determined by cluster-based permutation testing (*p < .05).

